# Towards a major methodological shift in depression research by assessing continuous scores of recurrence of illness, lifetime and current suicidal behaviors and phenome features: focus on atherogenicity and adverse childhood experiences

**DOI:** 10.1101/2023.02.26.23286462

**Authors:** Michael Maes, Ketsupar Jirakran, Asara Vasupanrajit, Patchaya Boonchaya-Anant, Chavit Tunvirachaisakul

**Affiliations:** Department of Psychiatry, Faculty of Medicine, Chulalongkorn University and King Chulalongkorn Memorial Hospital, the Thai Red Cross Society, Bangkok, Thailand; Cognitive Impairment and Dementia Research Unit, Faculty of Medicine, Chulalongkorn University, Bangkok, Thailand; IMPACT Strategic Research Center, Barwon Health, Geelong, Australia; Department of Psychiatry, Medical University of Plovdiv, Plovdiv, Bulgaria; Center of Excellence for Maximizing Children’s Developmental Potential, Department of Pediatric, Faculty of Medicine, Chulalongkorn University, Bangkok, Thailand; Division of Endocrinology and Metabolism, Faculty of Medicine, Chulalongkorn University, Bangkok, Thailand

**Keywords:** major depression, biomarkers, precision psychiatry, neuro-immune, inflammation, oxidative stress

## Abstract

**Background:** The binary major depressive disorder (MDD) diagnosis is inadequate and should never be used in research.

**Aims:** The study’s objective is to explicate our novel precision nomothetic strategy for constructing depression models based on adverse childhood experiences (ACEs), lifetime and current phenome, and biomarker (atherogenicity indices) scores.

**Methods:** This study assessed recurrence of illness (ROI: namely recurrence of depressive episodes and suicidal behaviors), lifetime and current suicidal behaviors and the phenome of depression, neuroticism, dysthymia, anxiety disorders, and lipid biomarkers (including ApoA, ApoB, free cholesterol and cholesteryl esters, triglycerides, high density lipoprotein cholesterol) in 67 normal controls and 66 MDD patients. We computed atherogenic and reverse cholesterol transport indices.

**Results:** We were able to extract one factor from a) the lifetime phenome of depression comprising ROI, and traits such as neuroticism, dysthymia and anxiety disorders, and b) the phenome of the acute phase (based on depression, anxiety and quality of life scores). PLS analysis showed that 55.7% of the variance in the lifetime + current phenome factor was explained by increased atherogenicity, neglect and sexual abuse, while atherogenicity partially mediated the effects of neglect. Cluster analysis generated a cluster of patients with major dysmood disorder, which was externally validated by increased atherogenicity and characterized by increased scores of all clinical features.

**Conclusions:** The outcome of depression should not be represented as a binary variable (MDD or not), but rather as multiple dimensional scores based on biomarkers, ROI, subclinical depression traits, and lifetime and current phenome scores including suicidal behaviors.

## Introduction

Recent research has demonstrated that binary major depressive disorder (MDD) diagnoses are inadequate and should never be used in clinical research [1, 2]. Binary diagnoses, such as for example, those proposed by the Diagnostic and Statistical Manual of Mental Disorders, Fifth Edition [3] and the International Classification of Disease [4], are mere obstacles to clinical depression research.

### Why the DSM and ICD diagnoses are impediments

First, the diagnosis of MDD is a post-hoc construction based on clinical narratives of the condition, including assessments of the current phenome (symptoms) and phenomenology (self-assessed disease features) and lifetime history [1,2,5,6]. The first question is why a post-hoc, binary construct should be used as the final outcome variable in research when continuous ratings that reflect current and lifetime deviations in the patient’s phenome and phenomenology can be assessed. It is well known that information is lost when continuous data is converted to binary data and that it is more difficult to achieve a higher level of precision and power with binary data. More unique assessments of the outcome variable (the phenome) mean more information, and the latter means more power and precision [1, 7].

Moreover, the binary diagnosis does not contain any information on important clinical outcome variables, such as suicidal behaviors. Furthermore, in clinical research, the diagnosis of MDD frequently combines responders to treatment + partial responders + remitted patients + non-responders or non-remitters, making it impossible to detect state MDD biomarkers. Another significant issue is that the lifetime severity of the diagnosis is not included in the MDD diagnosis.

Second, the binary diagnosis does not contain any information on the most important aspect of mood disorders, namely the recurrence of illness (ROI) index, which reflects the severity of recurrence of depressive episodes (and mania episodes in bipolar disorder), and recurrence of lifetime suicidal ideation (SI) and attempts (SA) [1,2,5,6,7]. Furthermore, one can diagnose single and recurrent depression using DSM [3] and ICD-11 [4] criteria, but these are again binary diagnoses. The ROI index is the most significant component of MDD in precision models because it is a continuous variable that predicts current SI and SA, the phenome and phenomenome of MDD, with high accuracy [1,5,6,7]. In addition, the ROI index is strongly associated with numerous biomarkers, such as genetic variants in Q192R paraoxoase 1 (PON1), PON1 activity, high density lipoprotein cholesterol (HDLc), lipid peroxidation, and immune activation [6,8,9,10,11].

Third, the DSM/ICD binary diagnoses do not allow to differentiate MDD from normal human stress-related responses including demoralization, feeling blue or sad, grief, and emotional distress [1,7,8]. A major problem in clinical MDD research is that these normal human responses to stress are lumped together with very severe phenotypes (including feelings of guilt, suicidal ideation, loss of insight, diurnal variation, anhedonia, hypoesthesia), thereby blurring the actual relationships between biomarkers and the actual state of severe depression phenotypes [1,5,7,8].

Fourth, using the binary diagnosis of MDD not only results in the loss of important information, but also in the use of a diagnosis with extremely low validity and reliability, including inter-rater reliability [1,2,5,7]. Even among psychiatrists, the use of the binary diagnosis of MDD results in an unacceptable number of misdiagnoses and misclassifications [1, 2]. Using such a diagnosis in research entails using an incorrect outcome model, whereas statistical analysis and machine learning (ML) techniques are only possible when a correct outcome model is used. The latter consists of three elements: model, correct, and outcome. a) Model means that the used model must meet Poppers’ [12] criteria and be confirmable, falsifiable, truth-value determinable, progressive, flexible, and adaptable. The DSM/ICD case definitions of MDD, on the other hand, do not meet any of the aforementioned criteria [1,2,5,7]. b) “Correct” denotes that the model must be statistically validated and cross-validated [1]. c) “Outcome” means that the “diagnostic model” should be conceptualized as the dependent variable in statistical or ML techniques [1,5,7]. Nonetheless, the incorrect and invalid binary diagnosis of MDD is most frequently used as an explanatory variable in analyses of variance or as a mediator variable in regression analyses. As a result, the majority of psychiatric research employs incorrect post-hoc, higher-order binary diagnoses in incorrect models (which transform dependent variables into explanatory variables), that are analyzed using improper statistical tests (inadequate t tests versus predictive models) [1,2,5]. Consequently, their use as an explanatory variable in statistical analyses is not only conceptually flawed, but also leads to an abundance of errors and incorrect conclusions [1,2,5,7].

### The only way forward: precision nomothetic psychiatry

The proper approach to defining a “correct outcome model” of depression research is to employ continuous variables reflecting a) the current phenome, comprising current suicidal behaviors (SBs), the severity of illness as assessed with depression interview-based ratings scales, and self-rated scales assessing depression, health-related quality of life (HR-QoL) and disabilities [1,2,5,7,10], and b) the lifetime phenome, which comprises ROI and different traits [13], including neuroticism, a form fruste of depression, dysthymia and anxiety disorders, such as generalized anxiety disorder (GAD) [5,7,13,14]. Genetic and trait biomarkers should consequently be used as predictors in ML techniques to predict the correct outcome model of depression, and state biomarkers should be used to examine associations with current phenome data.

However, such an approach cannot be implemented until the current academic norms to employ the gold standard DSM/ICD diagnosis and the psychiatrists’ tradition of binary thinking are abandoned to make room for a methodological transformation. New structures of reasoning, theoretical approaches, and methodologies are required to solve the mystery of depression and the role of its pathways. To accomplish this, we created precision nomothetic psychiatry, a new clinimetrics method based on (un)supervised ML (1,2,5,6,7,9-11]. The term “nomothetic” denotes that our method permits the extraction of mathematical laws or algorithms from a set of independent variables, including gene variants, early lifetime trauma, and adverse outcome pathways (AOPs), that explain the variability in the correct outcome models, consisting of the continuous ROI and phenome variables [1,2,5,7,9–11].

As previously described, this method allowed us to construct a) new pathway phenotypes, a combination of biomarkers and clinical phenome data into one validated construct, and b) new phenotype or endophenotype classes, which are groups of patients based on different features (e.g., a combination of ROI and phenome data) or phenotype and biomarker data, respectively [1,2,5,7,9–11]. For instance, we were able to establish a new phenotype class, namely major dysmood disorder (MDMD), which is distinguished from simple dysmood disorder (SDMD) by highly elevated ROI, LT, cognitive disorders, biomarkers, current SB, phenome and disability scores, and lowered HR-QoL [1,5,7]. In our previous models, however, we did not include depressive traits such as dysthymia, neuroticism, and anxiety disorder, nor did we examine whether ROI, these depressive traits, and the current phenome can be combined into a single dimension.

### Usefulness of the nomothetic approach in clinical practice: RADAR graphs

In addition, we devised a method for displaying all these distinct features of depression in a RADAR or spider graph, in which all the patient’s features are represented as scores and expressed as the distance in standard deviations from the controls (set to zero) [5, 7]. These scores, designated Research and Diagnostic Algorithmic Rules (RADAR), are comprised of ACEs, ROI, lifetime SBs, current phenome and SB scores, and the lifespan trajectory, which is a calculated composite of all patient characteristics, including ACEs. Consequently, the RADAR scores form a personalized RADAR graph that, like a fingerprint, authenticates a specific patient. This also demonstrates, from a clinical standpoint, how simplistic and minimal the binary diagnosis of MDD is in reality. In fact, our method uses many multiple bits instead of one incorrect bit (DSM/ICD), thereby increasing power, precision, and information. Nevertheless, until now we did not include biomarker RADAR scores in our spider graphs. This is significant because a RADAR score based on biomarkers may directly identifies the drug target to treat a specific patient in a personalized manner [5, 7].

### AOPs in depression precision nomothetic models

Previously, five distinct precision nomothetic models were built. **Figure 1** shows the theoretical frameworks that were used to develop these precision nomothetic models including causome variables (genetic variants, adverse childhood experiences), adverse outcome pathways (AOPs), lifetime and current phenome data such as symptoms and suicidal behaviors. A first nomothetic model was a clinical precision model that incorporated the effects of ACEs on ROI and the current phenome, such as physical and emotional abuse, physical and emotional neglect, and sexual abuse [1,5,6,7,11,15]. The second model included the effects of PON1 genetic variants, decreased PON1 enzymatic activity (gene-determined), decreased HDLc, and increased lipid peroxidation as trait and state biomarkers of ROI or the current phenome scores [9–11]. A third nomothetic model was constructed which linked elevated bacterial translocation with leaky gut and the subsequent autoimmune responses to lipid-associated neoepitopes (such as malondialdehyde) with ROI and the phenome [16]. A fourth model demonstrated that ROI and activation of the pro-inflammatory immune network could be combined with ROI, and that this new pathway-phenotype accurately predicted the current phenome of depression [6]. A fifth model incorporated gut enterotypes (a specific gut microbiota composition) in association with ROI and the phenome of depression [17].

**Figure 1.**
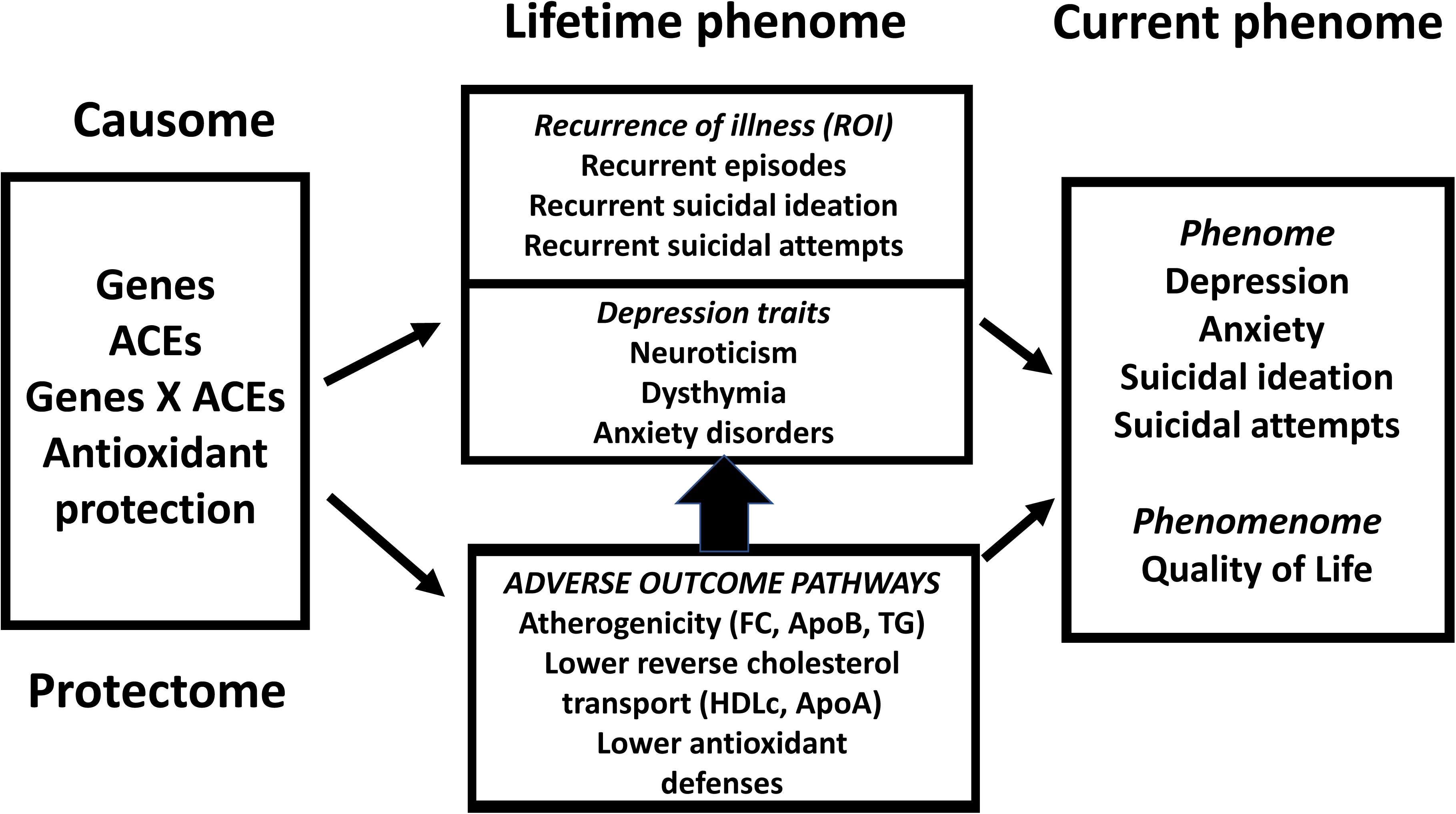
Theoretical framework of the precision nomothetic approach. ACE: adverse childhood experiences; FC: free cholesterol, Apo: apolipoprotein, TG: triglycerides, HDLc: high-density lipoprotein cholesterol.

To date, however, we have not developed precision nomothetic models that incorporate atherogenicity biomarkers. This is extremely important because decreased levels of the paraoxonase 1 (PON1) – high density lipoprotein cholesterol (HDLc) complex in serum (the PON1 enzyme is bound to HDLc) are strongly associated with the phenome of depression, including SBs [9-11,18], whereas MDD and neuroticism are strongly associated with elevated levels of free cholesterol (FC), triglycerides (TG), apolipoprotein (Apo)B, and ApoB/ApoA ratio, as well as decreased levels of HDLc and ApoA [13]. Interestingly, these results were only obtained after excluding all subjects with metabolic syndrome (MetS). The results suggest that MDD is distinguished by an increase in atherogenicity and a decrease in reverse cholesterol transport (RCT), a major antioxidant and protective pathway that protects against atherogenicity, lipid peroxidation, inflammation, and increased bacterial load [13]. Moreover, the increased atherogenicity in MDD also explains the strong comorbidity of MDD with atherosclerosis and cardiovascular disorders (CVD) [19–23].

#### Aims of the current study

The study’s overarching objective is to explicate our novel precision nomothetic strategy for constructing a depression model based on causome, AOP and lifetime and current phenome data. The specific aims are (a) to develop a precision nomothetic model of depression that links ACEs with ROI (recurrence of episodes and suicidal behaviors), depressive traits and the severity of the phenome of an acute phase of depression; b) to test whether ROI, subclinical traits and the severity of the phenome of an acute phase shape one dimension of depressive psychopathology; c) to construct a nomothetic model based on atherogenicity indices that mediate the effects of effects of ACEs on depressive traits, ROI and the phenome of depression; and d) to generate RADAR graphs which show the relevant LT and current phenome scores and atherogenicity indices in a fingerprint-like display.

## Methods and Participants

### Participants

There was a total of 133 people involved in this study: 66 people with major depression and 67 age- and sex-matched controls; 64 people with MetS and 69 people without it. Accordingly, we enrolled a total of 31 people with MDD and 34 controls, 35 patients with MDD who did not have MetS, 33 patients with MetS but not MDD, and 34 healthy individuals as comparisons. For this study, we invited both male and female Thai speaking people, aged 18-65, for participation. In the period beginning in September 2021 and continuing through February 2022, the Department of Psychiatry at King Chulalongkorn Memorial Hospital in Bangkok, Thailand, recruited outpatients and controls to be included in the study. Both the healthy controls and the MDD patients were recruited from the same geographical region, which was Bangkok in Thailand. The healthy controls included members of the personnel, members of the personnel’s families or acquaintances, and friends of MDD patients. Patients were given a diagnosis of MDD after being evaluated using the DSM-5 criteria [3]. Controls with a lifetime or current diagnosis of MDD were excluded from the study. We diagnosed MetS by using the 2009 Joint Scientific Statement published by the American Heart Association and the National Heart, Lung, and Blood Institute [24]. Patients with MetS and comorbid MDD+MetS were not excluded from the study when they were taking drugs for type 2 diabetes mellitus (T2DM), hypertension, obesity, dyslipidemia, atherogenicity, and diabetes, including glipizide, metformin, statins, amlodipine, losartan, liraglutide, fenofibrate, prazosin, and enalapril. Nonetheless, when using any of these medications, healthy controls and MDD patients in the study group who did not have MetS were excluded. Exclusion criteria for patients and controls included a) other axis-1 diagnoses beside MDD, including bipolar disorder, psycho-organic disorders, schizo-affective disorder, schizophrenia, eating disorders, autism, substance use disorders (SUDs) (except tobacco use disorder, TUD); and axis-2 diagnoses, such as borderline and antisocial personality disorder. Normal controls were also excluded if they had DSM-5 dysthymia or DSM-IV anxiety disorders such as GAD, post-traumatic stress disorder (PTSD), obsessive-compulsive disorder (OCD), agoraphobia, social phobia, or panic disorder. Other exclusion criteria for patients and controls were: a) pregnant and lactating women, b) immune and autoimmune disorders, such as chronic obstructive pulmonary disease, chronic kidney disease, systemic lupus erythematous, rheumatoid arthritis, psoriasis, inflammatory bowel disease, scleroderma, c) abnormal kidney and liver function tests, d) neurodegenerative or neuroinflammatory disorders, such as stroke, multiple sclerosis, epilepsy, Alzheimer and Parkinson disease, and brain tumors; e) use of therapeutic doses of antioxidant and omega-3 supplements, and f) current or lifetime use of immunomodulatory or immunosuppressants, including hydroxychloroquine, glucocorticoids, methotrexate, or pomalidomide.

All relevant national and international privacy and ethical guidelines were followed during the research. The study was approved by the Chulalongkorn University Medical Faculty’s Institutional Review Board (#445/63) in Bangkok, Thailand. Prior to participating in the study, all subjects were asked to voluntarily fill out a consent form with detailed information about their participation.

### Clinical measurements

This study’s socioeconomic and clinical data (including number of depressive episodes, medical and psychiatric history) were gathered using a semi-structured questionnaire given to both patients and controls. Mood disorders, suicide, or suicidal attempts (FHID_MDDSB) and SUDs (FHIS_SUD) in participants’ first-degree relatives were evaluated by a family history interview. We made the diagnosis of MDD by applying the Diagnostic and Statistical Manual of Mental Disorders, Fifth Edition (DSM-5) criteria [2] and the Mini International Neuropsychiatric Interview (M.I.N.I.) in a Thai translation [25]. The Columbia Suicide Severity Rating Scale (C-SSRS) was used for the evaluation of lifetime and current suicidal ideation or attempts [14, 26]. The ACE Questionnaire, in a Thai translation, was used to evaluate the presence of ACEs [27]. There are a total of 28 questions on this questionnaire, which cover a wide range of traumatic childhood experiences in 10 categories: emotional abuse (2 questions), physical abuse (2 questions), sexual abuse (4 questions), emotional neglect (5 questions), physical neglect (5 questions), domestic violence (4 questions), substance abuse in the home (2 questions), mental illness in the home (2 questions), parental divorce (1 question), and a criminal household member (1 item). Moreover, the M.I.N.I. was employed to make current and lifetime diagnoses of dysthymia, PTSD, GAD, panic disorder, OCD, social phobia, and agoraphobia. In addition, we diagnosed individuals with lifetime anxiety disorders using DSM-IV criteria and determined the diagnosis of dysthymia based on DSM-IV criteria. Neuroticism was measured using a Thai version of the Big Five Inventory [28, 29]. Since neuroticism is the sole factor consistently linked to MDD, we used raw scores from that dimension in the current study [14]. Both the Beck Depression Inventory II (BDI-II) [30] and the Hamilton Rating Scale for Depression [31] were used to assess depressive symptoms. The severity of anxiety was evaluated with the use of the State-Trait Anxiety Inventory (STAI), state version [32], which was translated into Thai by Iamsupasit and Phumivuthisarn [33]. We used the World Health Organization Quality of Life Instrument-Abbreviated Version (WHO- QoL-BREF) [34] to assess HR-QoL on the same day as the semistructured interview and rating scale scores. This measure assesses HR-QoL in four domains (physical health, mental health, social interactions, and the environment) across a total of 26 items. Using the WHO-QoL-BREF criteria [34], we determined the unweighted raw scores for the four domains.

The American Heart Association and the National Heart, Lung, and Blood Institute published a joint scientific statement [24] defining MetS as the presence of three or more of the following components: (1) having a waist circumference of at least 90 centimetres for men and at least 80 centimetres for women, or having a body mass index of at least 25 kilogrammes per square metre; (2) having a high triglyceride level of at least 150 milligrammes per deciliter; (3) having a low HDL cholesterol level of at least 40 milligrammes per deciliter for men and at least 50 milligrammes per deciliter for women; (4) having high blood pressure, namely 130 mm Hg systolic blood pressure, or 85 mm Hg diastolic blood pressure or treatment with antihypertensive medication; and (5) high fasting blood glucose 100 milligrammes per deciliter or having a diabetes diagnosis. Body mass index (BMI) is calculated by dividing weight (kg) by height (m2) squared.

### Assays

In order to analyse lipid biomarkers, blood was collected at 8:00 a.m. after an overnight fast and no more than 48 hours after hospital admission. Frozen serum samples were stored at 80 °C until analysis. The Alinity C was used to determine TC, HDLc, TG, and direct LDL, as previously described [13] (Abbott Laboratories, USA; Otawara-Shi, Tochigi-Ken, Japan). Coefficients of variance for TG, HDLc, TG, and LDLc were 2.3%, 2.6%, 2.3%, and 4.5%, respectively. Immunoturbidimetric assays using the Roche Cobas 6000 and c501 module were used to determine levels of Apo A1 and Apo B. (Roche, Rotkreuz, Switzerland). Apo A1 and Apo B had intra-assay CV values of 1.75% and 2.64%, respectively. The Free Cholesterol Colorimetric Assay Kit was used to measure the amount of free cholesterol (FC) (Elabscience, cat number: E-BC-K004-M).

### Statistical analysis

In order to compare nominal variables across categories, either the Chi-square test or Fisher’s exact probability test was utilized. To examine the differences in scale variables between the control group and the MDD patients, an analysis of variance (ANOVA) was conducted. Correlations between pairs of scale variables were determined using Pearson’s product-moment correlation coefficients. The most important biomarkers predicting clinical data (such as lifetime phenome) were delineated using multiple regression analysis (a manual approach). In addition, a forward stepwise automated regression approach with p-values of 0.05 to-enter and 0.1 to delete was used to determine the best predictors of the model. In the final regression models, we calculated the standardized β coefficients using t-statistics and exact p-values for each explanatory variable in addition to the model’s F statistics (and p values) and total variance (R^2^ or partial eta squared as effect size). The White and the modified Breusch-Pagan was utilized to check the existence of heteroskedasticity. Tolerance (cut-off value 0.25) and variance inflation factor (cut-off value > 4) as well as the condition index and variance proportions from the collinearity diagnostics table were utilized to assess the presence of collinearity and multicollinearity. The calculated partial regression graphs showed the independent relationships between the dependent and explanatory variables. We considered an alpha value of 0.05 to be statistically significant for every one of the above tests. Only when the Kaiser-Meyer-Olkin test for factorability (satisfactory when > 0.5 and adequate when > 0.7), Bartlett’s sphericity test, anti-image correlation matrix, variance explained by the first PC (>50%), and all loadings on the first PC are all > 0.65, then the first PC is accepted as a validated construct [5, 7]. IBM’s SPSS for Windows, Version 28 was used in this study.

Partial Least Squares (PLS) Analysis was used to build the nomothetic models, and we provided a summary of this method in previous papers [1,5,6,7,10,15]. Previously, we described the Heterotrait-Monotrait (HTMT) ratio [35] and the Invariance Assessment of Composite Models (MICOM) [7], two additional, crucial PLS tests for evaluating the discriminant validity of the constructs and invariance assessments, respectively.

## Results

### Construction of ACE causome factors

We [14] used principal component analysis (PCA) to determine three important ACE categories: a) using ACE9, ACE10, ACE11, ACE12, ACE13, and ACE15 (physical and emotional neglect items, for a full list and description of the ACE components, see Table 1 in the Electronic Supplementary File in [14]), we created a PC that we named “ACEneglect”; b) using ACE1, ACE2, ACE3, ACE4, ACE19, ACE20, ACE21, and ACE22 (all physical and emotional abuse ACE items), we derived a second validated factor, named “ACEabuse” [14]; and c) using ACE5, ACE6, ACE7, and ACE8 (all sexual abuse items) we constructed a third PC, labeled ACEsexabuse [14]. The other ACE rating scale items were not significant in the current analysis.

**Table 1.** Correlations between adverse childhood experiences (ACEs), recurrence of illness (ROI) and the phenome and phenomenome of depression Results of Pearson correlation calculations (all n=133). ROI: recurrence of illness, a principal componenet (PC) based on lifetime suicidal behaviors (SB) and recurrence of episodes; LT_phenome: PC extracted from ROI, neuroticism, dysthymia, and lifetime anxiety disorders; Current_phenome: the first PC extracted from current scores on Beck Depression Inventory; Hamilton Depression Rating Scale, State and Trait Anxiety Inventory, current_SBs, and the first PC extracted from the health-related quality of life (HR-QoL) measurements.

| Variables | ACE neglect | ACE abuse | ACE sexual abuse | ROI |
| --- | --- | --- | --- | --- |
| ROI | 0.443 (<0.001) | 0.300 (<0.001) | 0.204 (0.019) | - |
| LT_phenome | 0.425 (<0.001) | 0.334 (<0.001) | 0.226 (0.009) | - |
| Current_phenome | 0.460 (<0.001) | 0.298 (<0.001) | 0.374 (<0.001) | 0.741 (<0.001) |
| LT+Current_phenome | 0.464 (<0.001) | 0.373 (<0.001) | 0.264 (<0.001) | - |
| HR-QoL_physical | -0.526 (<0.001) | -0.334 (<0.001) | -0.233 (0.007) | -0.514 (<0.001) |
| HR-QoL_mental health | -0.498 (<0.001) | -0.342 (<0.001) | -0.232 (0.007) | -0.509 (<0.001) |
| HR-QoL_social | -0.453 (<0.001) | -0.245 (<0.004) | -0.142 (0.104) | -0.427 (<0.001) |
| HR-QoL_environment | -0.374 (<0.001) | -0.224 (0.010) | -0.157 (0.070) | -0.295 (<0.001) |

### Construction of adverse outcome pathways

To determine the cholesterol esterification rate (CER), we use the following formula: 1 – (free cholesterol / total cholesterol) x 100 [20]. The ApoB/ApoA ratio and the AIP index (TG / HDLc) were calculated. Three novel indices were calculated [13]: a) a pro-atherogenicity index as z transformation of ApoB (z ApoB) + z TG + z LDLc + z FC (dubbed PRO_AI); b) an anti-atherogenic potential (ANTI_AI) also reflecting RCT as: z HDLc + z ApoA + z CER; and c) their ratio computed as z PRO_AI – z ANTI_AI, dubbed as PRO/ANTI_AI [13].

### Construction of the lifetime phenome of depression

The number of major depressive episodes (self-reported by the patient and outpatient clinical records) and SBs in one’s lifetime formed the basis for the ROI index. For the reasons stated above, we calculated three indices of SBs over the course of a person’s lifetime until one month before the current episode [5, 14]. Ten LT SI items (AVE=0.741, Cronbach alpha=0.961, all loadings > 0.803) and 5 LT SA items (AVE=0.566, Cronbach alpha=0.809, all loadings > 0.657) each contributed to a single latent vector, which represents LT_SI and LT_SA, respectively [14]. As a result, we created a single composite indicating LT_suicidal behaviors (designated LT_SBs) that is a z-unit weighted composite based on LT_SI and LT_SA scores [14].

**Figure 2** shows a first PLS model which demonstrates how we constructed ROI, a trait score (labeled LT_traits), an index of the lifetime phenome (LT_phenome). ROI was conceptualized as the first factor extracted from LT_SI and LT_SA, number of depressive episodes as self-reported by the patient and registered in the outpatient records (OPD) (average variance extracted or AVE=64.5%; Cronbach’s alpha=0.817; composite reliability=0.879, rho_A=0.829; all loadings > 0.746). PLS Blindfolding shows that the construct cross-validated redundancies were significant (this is the case for all following PLS constructs described in the paper). Furthermore, this figure shows that one latent vector could be extracted from neuroticism scores, lifetime anxiety disorders, and dysthymia (AVE=62.7%; Cronbach’s alpha=0.704; composite reliability=0.834, rho_A=0.715).

**Figure 2.**
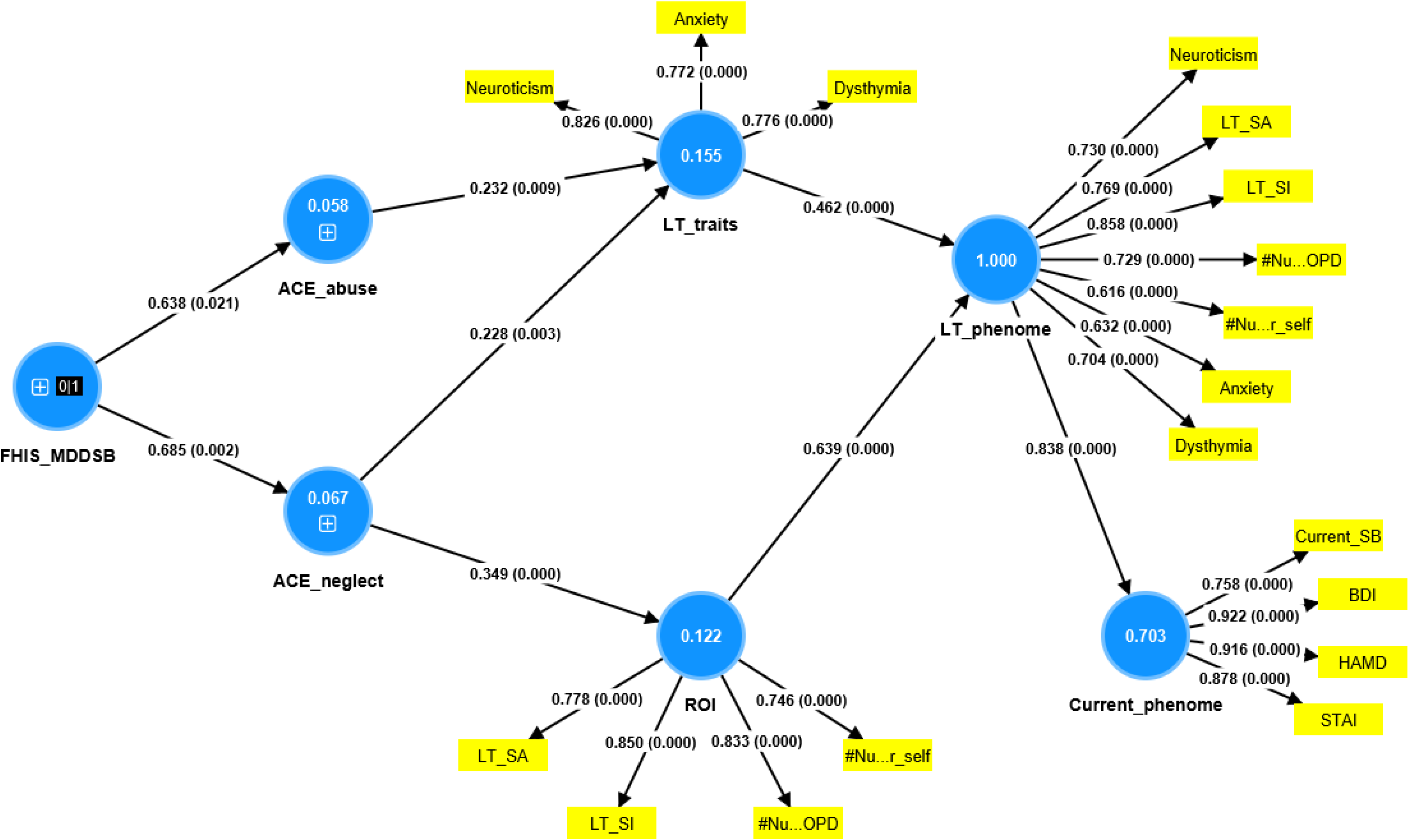
A first partial least squares (PLS) model. This model shows the construction of the recurrence of illness (ROI) index, a trait depression score (LT_traits), an index of the lifetime phenome (LT_phenome), as well as the severity of the current phenome (Current_phenome). LT_SI: lifetime suicidal ideation; LT_SA: lifetime suicidal attempts; #nu ..OPD and #nu..r_self: number of depressive episodes as registered in the outpatient records (OPD) and self-reported by the patient, respectively. LT_phenome: is a higher-order construct based on all ROI and LT_traits indicators. Current_phenome: is a factor extracted from current suicidal behaviors, and scores on the Beck Depression Inventory (BDI), Hamilton Depression Rating Scale (HAMD), State and Trait Anxiety Inventory (STAI).

### Construction of the current phenome of depression

Seven items from the C-SSRS were used to derive the first PC reflecting current (last month) suicidal ideation (Current_SI, KMO=0.703, Bartlett’s χ^2^=982.510, df=15, p<0.001, EV=72.05%, all loadings > 0.672; [14]). The severity of current SA (Current_SA) was calculated using the first PC extracted from 5 C-SSRS items [14] (KMO=0.636, Bartlett’s χ2=302.236, df=3, p<0.001, EV=81.14%, all loadings > 0.807). Current SBs (Current_SBs) were conceived of as a z-unit-weighted composite of Current_SI and Current_SA. Figure 2 shows that we were able to extract one factor from HAMD, STAI, BDI, and Current_SBs (AVE=75.9%; Cronbach’s alpha=0.891; composite reliability=0.926, rho_A=0.892; all loadings > 0.758). As such, we constructed a latent vector reflecting the severity of the current phenome labeled Current_phenome 1.

### Socio-demographic and clinical data in MDD and controls

ESF, Table 1 shows that there were no significant differences between MDD and controls in socio-demographic data except in the unemployment ratio. Normal controls and MDD patients without MetS were free of any medical drugs, including antihypertensive and antidiabetic drugs, etc. Part of the patients were taking psychotropic drugs, including sertraline (n=28), trazodone (n=16), fluoxetine (n=7), venlafaxine (n=13), paroxetine (n=2) and agomelatine (n=2) mirtazapine (n=8), benzodiazepines (n=36) and mood stabilizers (n=2), but these drugs did not have any effects on the biomarkers as shown previously using the same data set [13]. ESF, Table 2 show the differences in lipid profiles between MDD and controls (patients with MetS excluded).

**Table 2.** Correlations between lipid composite indices, and adverse childhood experiences (ACEs), recurrence of illness (ROI) and the phenome and phenomenome of depression Results of Pearson correlation calculations (all n=69 after excluding all subjects with metabolic syndrome). ROI: recurrence of illness, a principal componenet (PC) based on lifetime (LT) suicidal behaviors (SB) and recurrence of episodes; LT_phenome: PC extracted from ROI, neuroticism, dysthymia, and lifetime anxiety disorders; Current_phenome 1: the first PC extracted from current scores on Beck Depression Inventory; Hamilton Depression Rating Scale, State and Trait Anxiety Inventory, and Current_SBs; Current_QoL: the first PC extracted from the health-related quality of life subdomains. Apo: apolipoprotein, AIP: atherogenic index of plasma, PRO_AI: an atherogenic index based on free cholesterol, triglycerides, ApoB, low- density lipoprotein cholesterol, RCT: an index of reverse cholesterol transport based on low-density lipoprotein cholesterol, Apo A and cholesterol esterification ratio.

| Variables | ApoB/ApoA | AIP | PRO_AI | RCT | PRO/ANTI_AI |
| --- | --- | --- | --- | --- | --- |
| ACE neglect | 0.208 (0.086) | 0.216 (0.075) | 0.072 (0.554) | -0.329 (0.006) | 0.297 (0.013) |
| ACE abuse | 0.158 (0.193) | 0.142 (0.245) | 0.112 (0.358) | -0.115 (0.346) | 0.165 (0.176) |
| ACE sexual abuse | 0.263 (0.029) | 0.077 (0.528) | 0.091 (0.455) | -0.110 (0.368) | 0.168 (0.168) |
| ROI | 0.360 (0.003) | 0.406 (<0.001) | 0.312 (0.010) | -0.245 (0.044) | 0.357 (0.003) |
| LT_phenome | 0.430 (<0.001) | 0.501 (<0.001) | 0.338 (0.005) | -0.324 (0.007) | 0.430 (<0.001) |
| Current_phenome 1 | 0.401 (<0.001) | 0.472 (<0.001) | 0.186 (0.126) | -0.343 (0.004) | 0.383 (<0.001) |
| LT+Current_phenome | 0.461 (<0.001) | 0.500 (<0.001) | 0.303 (0.012) | -0.354 (0.003) | 0.450 (<0.001) |
| Current_QoL | -0.265 (0.028) | -0.337 (0.005) | -0.029 (0.811) | 0.319 (0.007) | -0.277 (0.021) |

### Construction of a higher-order lifetime phenome model

HTMT analysis showed that no discriminant validity could be established between ROI and LT_traits (HTMT index=1.118). Therefore, Figure 2 examines whether all ROI and LT_traits indicators could be combined into one higher-order construct, namely LT_phenome consisting of ROI and LT_traits. One validated construct could be extracted from these 7 indicators (AVE=52.4%; Cronbach’s alpha=0.846; composite reliability=0.884, rho_A=0.859, all loadings > 0.616). As such, we were able to construct a higher-order construct reflecting the LT_phenome of depression. PLS pathway analysis (5,000 bootstraps) showed that 70.3% of the variance in the Current_phenome 1 was explained by LT_phenome. Importantly, 15.5% of the variance in LT_traits was explained by ACEabuse and ACEneglect, while 12.2% of the variance in ROI was explained by ACEneglect. Both ACE indicators were in part predicted by FHIS_MDDSB.

### Construction of a higher-order current phenome model

ESF, Figure 1 shows that one validated vector could be extracted from the 4 HR-QoL domains, labeled Current_QoL (AVE=70.8%; Cronbach’s alpha=0.863; composite reliability=0.906, rho_A=0.887; all loadings > 0.783). We found that 62.4% of the variance in Current_QoL was explained by Current_phenome 1. Nevertheless, no discriminant validity could be established between Current_phenome 1 and Current_QoL (HTMT index=0.864). ESF, Figure 2 shows that one latent vector could be extracted from Current_SBs, Current_phenome, and Current_QoL (labeled Current_phenome 2) (AVE=71.5%; Cronbach’s alpha=0.794; composite reliability=0.881, rho_A=0.827; all loadings > 0.857).

### Association between LT_phenome and Current_phenome

To examine the associations among LT_ and Current_phenome, we have performed another PLS analysis (ESF, Figure 2) with as dependent variable the first factor extracted from Current_SBs, Current_phenome, and Current_QoL (labeled Current_phenome 2), and as input variable a factor extracted from the LT_phenome manifestations (number of episodes entered as a composite based on both self-declared and OPD assessments and LT_SBs). We found that 66.4% of the variance in Current_phenome 2 was explained by LT_phenome. Nevertheless, no discriminant validity could be established because the HTMT ratio is as high as 0.985. Therefore, in the next step we examined whether LT_phenome and Current_phenome features could be combined into one factor. **Figure 3** shows that we were able to extract one factor from ROI, LT_traits, Current_QoL, Current_SBs, and Current_phenome 1, labeled LT+Current_phenome (AVE=68.3%; Cronbach’s alpha=0.881; composite reliability=0.914, rho_A=0.887; all loadings > 0.716).

**Figure 3.**
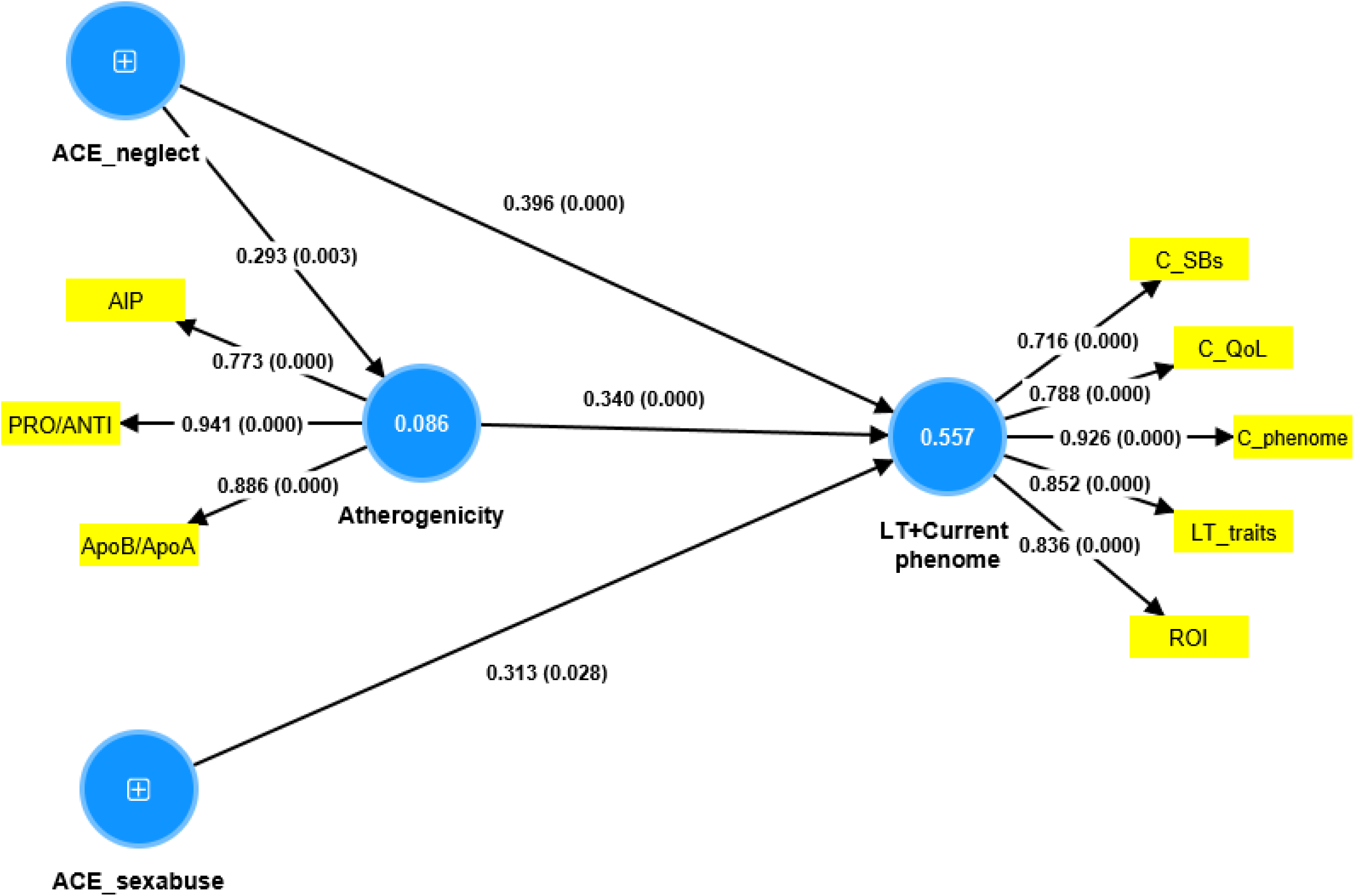
The final Partial Least Squares (PLS) model after feature reduction. This model shows the construction of the LT+Current_phenome conceptualized as a factor extracted from recurrence of illness (ROI), a trait score (LT_traits), the severity of the current phenome (C_phenome), a factor extracted from the 4 health-related quality of life scores (inversed C_QoL), and current suicidal behaviors (C_SBs) Atherogenicity was conceptualized as a factor extracted from the atherogenetic index of plasma (AIP), the apolipoprotein B (ApoB)/ApoA ratio, and a comprehensive pro-atherogenic / anti-atherogenic index (PRO/ANTI_AI). ACE: adverse childhood experiences.

### Correlations between clinical scores, ACEs and atherogenicity

Before performing a PLS analyses, it is always best to first examine the associations between the variables using correlation or regression analyses (see **Tables 1-3**). We found that ACEs, especially neglect and abuse, and ROI were strongly associated with the various clinical constructs and with the HR-QoL domains (Table 1). The regression and PLS analyses considering the effects of lipids were performed on subjects without MetS because (as mentioned in the Introduction) inclusion of subjects with MetS blurs the actual associations between lipids and psychopathology [13]. Table 2 shows the correlations between ACEs, ROI and clinical assessments and the lipid profiles. ACE neglect was significantly correlated with RCT and the PRO/ANTI_AI. ROI and the clinical phenome data were associated with most lipid profiles. All lipid indices containing anti-atherogenic assays were associated with Current_QoL.

**Table 3** shows the results of 4 regression analyses with the clinical outcome data as dependent variables. Regression #1 shows that 49.8% of the variance in the LT+Current_phenome was explained by the combined effects of ACE neglect and sexual abuse, PRO/ANTI_AI (all positively), and age (inversely). LT+Current_SBs (regression #2) were strongly (48%) predicted by ACEsexabuse and ACEneglect, PRO/ANTI_AI and age (inversely). Current_phenome 1 (regression #3) was predicted by ACEsexabuse and neglect and PRO/ANTI_AI. Regression #4 shows that 45.6% of the variance in Current_SBs was explained by ACEsexabuse and PRO/ANTI_AI.

**Table 3.** Results of multiple regression analyses with clinical data as dependent variables and biomarkers as explanatory variables LT+Current_phenome: PC extracted from recurrence of illness and lifetime suicidal behaviors (SBs), neuroticism, dysthymia, lifetime anxiety disorders, Beck Depression Inventory; Hamilton Depression Rating Scale, State and Trait Anxiety Inventory, current SBs; Current_phenome 1: PC extracted from Beck Depression Inventory; Hamilton Depression Rating Scale, State and Trait Anxiety Inventory, and current SBs, Current_suicidal behaviors: PC extracted from suicidal ideation and attempt items, ACE: adverse childhood experiences, Apo: apolipoprotein, PRO/ANTI_AI: a pro-atherogenic/antiantherogenic index, PRO: computed as a composite based on free cholesterol, triglycerides, ApoB, low-density lipoprotein cholesterol, ANTI: an index of reverse cholesterol transport based on low-density lipoprotein cholesterol, Apo A and cholesterol esterification ratio

| Dependent variables | Explanatory variables | Parameter estimates + statistics |  |  | Model statistics + effect sizes |  |  |  |
| --- | --- | --- | --- | --- | --- | --- | --- | --- |
|  |  | B | t | p | F<sub>model</sub> | df | p | R<sup>2</sup> |
| #1. LT+Current_phenome | **Model** |  |  |  | **15.62** | **4/63** | **<0.001** | **0.498** |
|  | ACE neglect | 0.330 | 3.42 | 0.001 |  |  |  |  |
|  | PRO/ANTI_AI | 0.302 | 3.24 | 0.002 |  |  |  |  |
|  | ACE sexual abuse | 0.251 | 2.67 | 0.010 |  |  |  |  |
|  | Age | -0.226 | -2.45 | 0.017 |  |  |  |  |
| #2. LT+Current suicidal behaviors | **Model** |  |  |  | **14.55** | **4/63** | **<0.001** | **0.480** |
|  | ACE sexual abuse | 0.392 | 4.11 | <0.001 |  |  |  |  |
|  | PRO/ANTI_AI | 0.286 | 3.03 | 0.004 |  |  |  |  |
|  | Age | -0.218 | -2.32 | 0.024 |  |  |  |  |
|  | ACE neglect | 0.215 | 2.18 | 0.033 |  |  |  |  |
| #3. Current_phenome 1 | **Model** |  |  |  | **26.17** | **3/65** | **<0.001** | **0.547** |
|  | ACE sexual abuse | 0.476 | 5.45 | <0.001 |  |  |  |  |
|  | ACE neglect | 0.340 | 3.77 | <0.001 |  |  |  |  |
|  | PRO/ANTI_AI | 0.203 | 2.31 | 0.024 |  |  |  |  |
| #4. Current_suicidal behaviors | **Model** |  |  |  | **27.72** | **2/66** | **<0.001** | **0.456** |
|  | ACE sexual abuse | 0.600 | 6.55 | <0.001 |  |  |  |  |
|  | PRO/ANTI_AI | 0.243 | 2.65 | 0.010 |  |  |  |  |

### Construction of the final PLS model based on ACE, atherogenicity and LT+Current_phenome

In Figure 3, we entered a factor (labeled atherogenicity LV) extracted from three major atherogenicity indexes using AIP, ApoA/ApoB and PRO/ANTI_AI (AVE=75.6%; Cronbach’s alpha=0.835; composite reliability=0.902, rho_A=0.833; all loadings > 0.716) as a direct predictor of LT+Current_phenome, while ACEs could predict LT+Current_phenome and the atherogenicity latent vector. The model standardized root mean squared residual (SRMR) value was 0.058, indicating an adequate model fit. PLS Predict shows that all Q2 predict values were positive and PLS Blindfolding shows that the construct cross-validated redundancies were significant. We found that 55.7% of the variance in LT+Current_phenome was explained by the atherogenicity LV, ACEneglect and ACEsexabuse, and that 8.6% of the variance in the atherogenicity LV was explained by ACEneglect, indicating that atherogenicity mediates part of the effects of ACEneglect on the phenome. PLS

It is interesting to note that we were able to extract one factor from the three LT_traits indicators, and the major atherogenicity indicators (AIP, ApoB/ApoA ratio, and PRO/ANRI_AI) (AVE=52.2%; Cronbach’s alpha=0.784; composite reliability=0.874, rho_A=0.812; all loadings > 0.676). As such, we have constructed an atherogenicity-traits pathway-phenotype, which explained 54.9% of the variance in Current_phenome 2. It should be mentioned that the lipid data could not be combined into one validated factor with Current_phenome 2.

### Construction of MDMD and SDMD classes

In order to construct classes within the MDD group, we performed two-step cluster analysis with the diagnosis MDD versus controls as categorical variable and 5 continuous variables, namely number depressive episodes, LT_SBs, LT_traits, Current_phenome (HAMD, STAI and BDI-II), and Current_SBs. Three clusters were formed with an adequate silhouette measure of cohesion and separation of 0.67, namely cluster 1 (n=67 controls), cluster 2 with less severe MDD features (n=30) and one with severe MDD features (n=35) as shown in **Figure 4**. There were no significant differences in any of the socio-demographic data between both groups, except the unemployment rate (higher in patients). All features listed were significantly different (p<0.01) between the three groups, except ACEabuse (no difference between cluster 1 and 2, p=0.132), ACEsexabuse (no difference between cluster 2 and 3, p=0.907), LT_SBs (no difference between cluster 1 and 2, p=0.263) and LT+Current_SBs (no difference among clusters 1 and 2, p=0.515).

**Figure 4.**
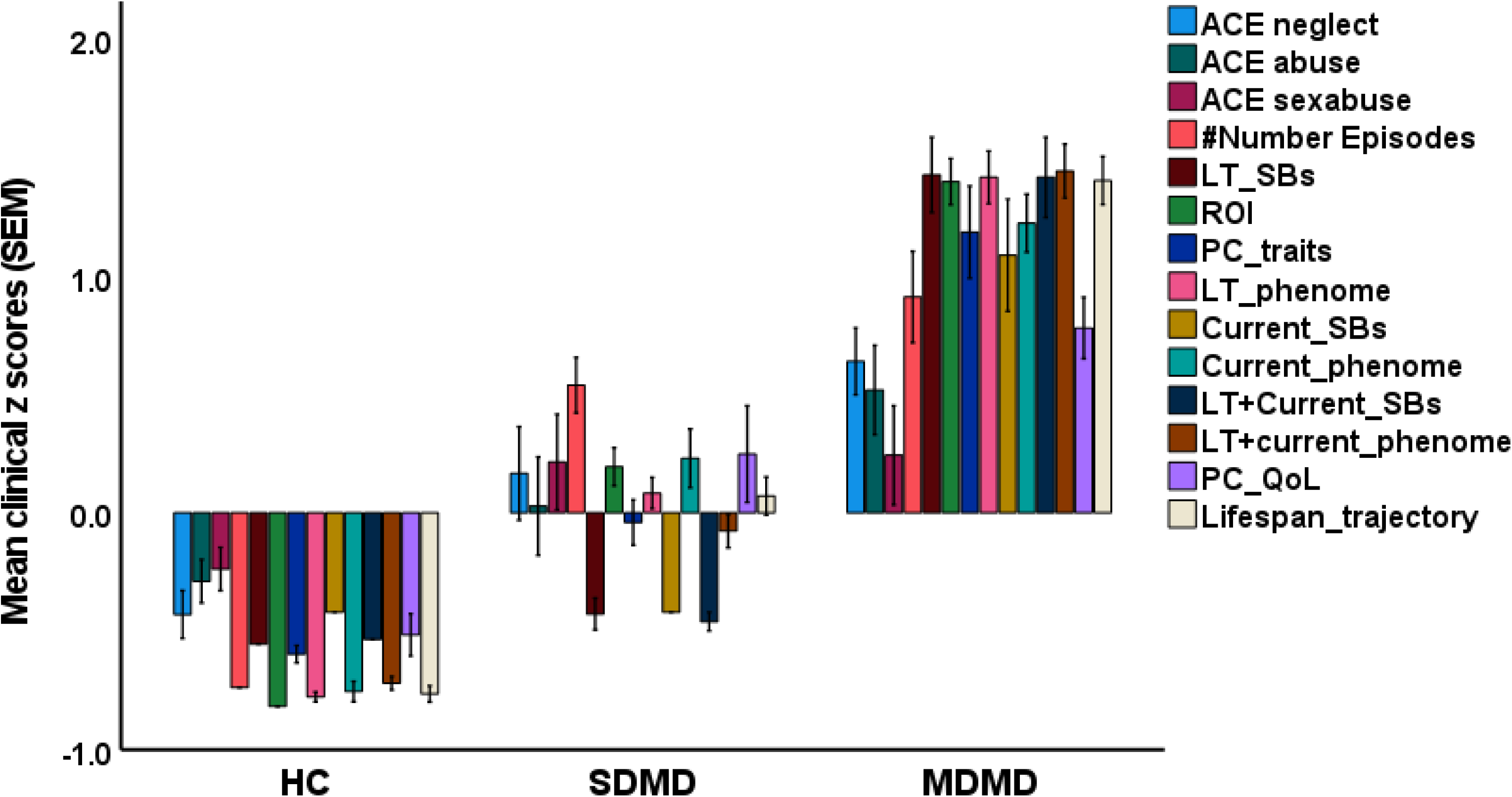
Results of cluster analysis with the formation of three clusters: healthy controls (HC), a cluster with severe clinical features, labelled major dysmood disorder (MDMD), and one with mild features, dubbed simple DMD (SDMD). ACE: adverse childhood experiences; LT_SBs: lifetime suicidal behaviors; ROI: recurrence of illness, a principal component (PC) based on LT_SBs and recurrence of episodes; LT_phenome: PC extracted from ROI, neuroticism, dysthymia, and lifetime anxiety disorders; Current_phenome: the first PC extracted from current scores on Beck Depression Inventory; the Hamilton Depression Rating Scale, State and Trait Anxiety Inventory, and current SBs; LT+Current_phenome: lifetime and current phenome; PC_QoL: the first PC extracted from the health-related quality of life subdomains (inverse values); Lifespan_trajectory: composite based on ACEs and all LT and current clinical variables.

### External validation of the MDMD class

**Figure 5** shows a bar graph with the relevant lipid profile measures in controls, SMDM and MDMD (after excluding all patients with MetS). We found significant increases in FC (p=0.044), TG (p=0.002), LDLc (p=0.040), ApoB (p=0.007), AIP (p<0.001), PRO_AI (p=0.004), and PRO/ANTI_AI (p<0.001) and lower HDLc (p=0.035) and RCT (p=0.030) in MDMD as compared with controls, whereas no significant changes could be established between SDMD and controls and between MDMD and SDMD. As such, the clinical, cluster analysis derived MDMD class was externally validated by lipid profiles.

**Figure 5.**
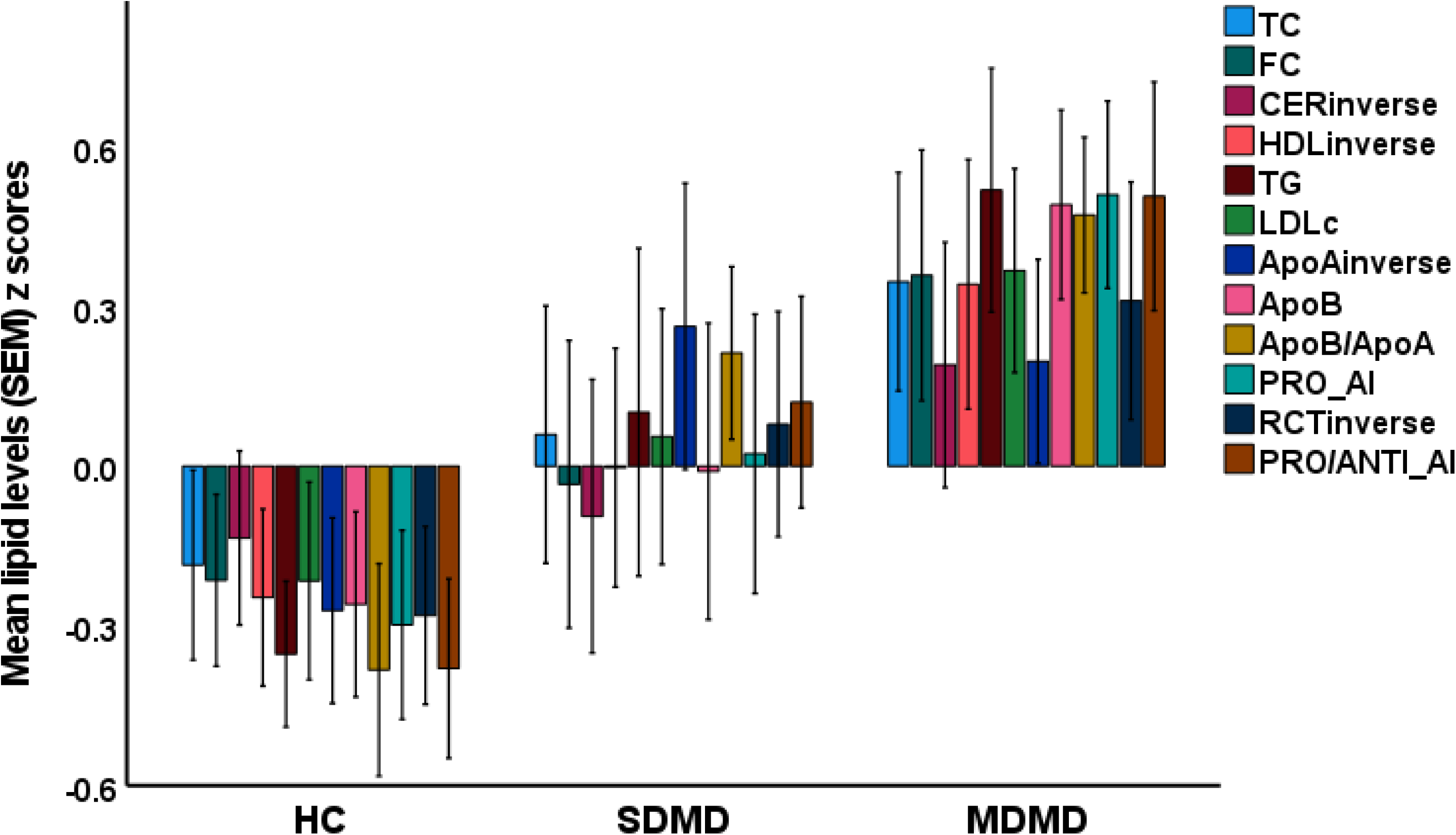
Lipid profiles of the cluster analysis-derived groups of healthy controls (HC), major dysmood disorder (MDMD), and simple DMD (SDMD). TC: total cholesterol, FC: free cholesterol, CER: cholesterol esterification rate, HDL: high density lipoprotein cholesterol, TG: triglycerides, LDL: low density lipoprotein cholesterol, Apo: Apolipoprotein, PRO_AI: pro-atherogenic index based on FC, ApoB, TG, and LDL; RCT: reverse cholesterol transport index based on HDL, ApoA and CER, PRO/ANTI_AI: PRO/AI/RCT index. Some data are shown in inverse transformation (CER, HDL, ApoA, and RCT).

### Construction of RADAR graphs

**Figure 6** depicts the RADAR scores for two patients, one with MDMD and another with SDMD. The RADAR graph displays fifteen RADAR scores on fifteen radial axes, each of which corresponds to a feature of mood disorders. The features are ordered according to the individual’s lifespan trajectory, beginning with ACEs, then LT clinical features (ROI and traits), current features (phenome, SBs, and HR-QoL) and a lifespan trajectory score, which was a z unit-weighted composite based on all features. The common centre point (zero score) was established as the mean value of all RADAR scores of the healthy controls set to 0. The RADAR score of the patients is expressed in z scores adjusted to the mean (set at 0) of healthy controls and thus displays the difference in standard deviations between the scores of both patients and controls. Phrased differently, the RADAR graph displays the relative position of the patients’ feature scores as compared with controls and this allows one to evaluate the differences in RADAR chart area shapes between the patients (or means in diagnostic groups). In addition, Figure 6 demonstrates that the radial axes are joined in the middle of the figure (zero feature ratings for the normal controls) and are joined by angular axes that divide the graph into grids displaying the difference in feature ratings between the subjects and normal controls. This RADAR graph demonstrates that the RADAR charts of the two patients are vastly dissimilar, especially the atherogenicity indices, ROI, LT_traits, SB, phenome scores, and the lifespan trajectory scores.

**Figure 6.**
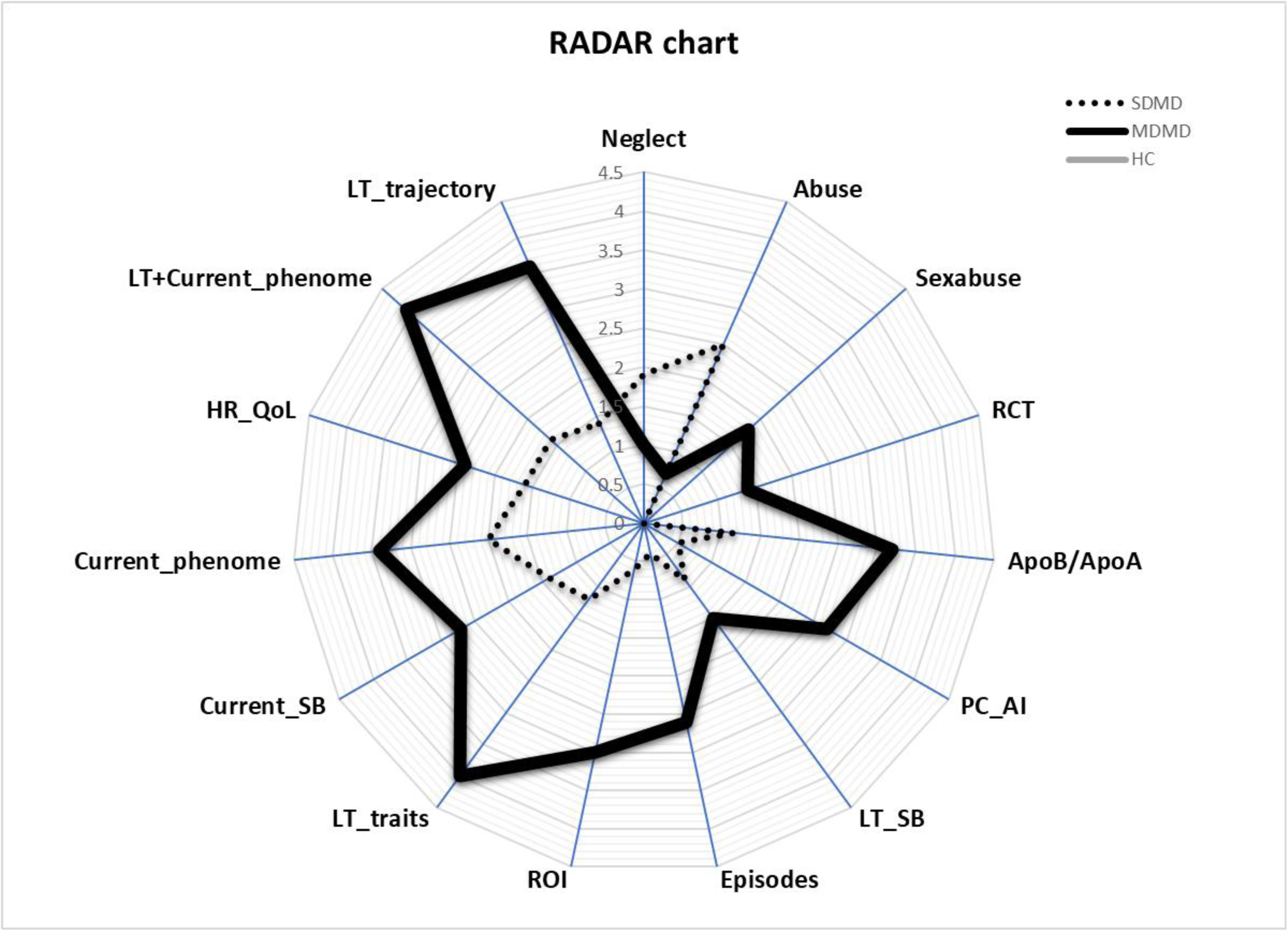
RADAR graph showing Research and Diagnostic Algorithmic Rule (RADAR) scores of two patients, one with simple dysmood disorder (SDMD) and one with major dysmood disorder (MDMD). The common centre point of this graph corresponds to the mean RADAR scores of the healthy controls (HCs), which were all set at 0. The RADAR scores are expressed as the difference from the centre point in standard deviations (SDs). The axes are interconnected, dividing the RADAR graph into grids that allow one to evaluate the difference in SD between patients and controls. ACE: adverse childhood experiences (physical and emotional neglect and abuse), Sex abuse: adverse childhood experiences, sexual abuse, RCT: reverse cholesterol transport, Apo: apolipoprotein, PC_AI: latent vector extracted from the pro-atherogenic / anti-atherogenic index, ApoB/ApoA and the ratio of triglyceride/high density lipoprotein cholesterol, LT_SB: lifetime suicidal behavior, Episodes: number of episodes, ROI: recurrence of illness, LT_traits: latent vector extracted from neuroticism, dysthymia, and anxiety disorders, Current_SBs: current suicidal behaviors, Current_phenome: latent vector extracted from depression and anxiety severity ratings and Current_SBs, HR_QoL: health-related quality of life, LT+Current_phenome: latent vector extracted from ROI, LT_traits, Current_phenome, and HR-Qol; LT_trajectory: lifespan trajectory.

## Discussion

### New clinical precision models of depression

In this research, we detailed the process of creating validated and replicable precision nomothetic models of depression. Using ACEs, ROI, depressive traits, lifetime and current SI and SA, and the acute phase phenome and phenomenome, we first tested various models and ultimately constructed a parsimonious final model after feature reduction. This new precision nomothetic model builds on earlier work [5,6,11,15,16,17] by integrating measures of atherogenicity and depressive traits, such as neuroticism, dysthymia, and anxiety disorders. We demonstrated how to calculate RADAR scores, which monitor the various aspects of the disorder, such as a) the severity of the acute phenome of depression, comprising symptom domains based on interviews (phenome) and self-rated domains, including HR-QoL (phenomenome), b) the lifetime phenome, comprising ROI (recurrence of episodes and SBs) and depression traits (neuroticism, dysthymia, and anxiety disorders); and c) atherogenicity. When compared to the unreliable DSM and ICD case definitions, still the gold standards in clinical practice and research, our nomothetic model represents a significant improvement. The benefit of our model can be summed up in one question: “why would someone stick with a false and unreliable single bit of information (DSM or ICD diagnosis) when numerous bits of information should be monitored across multiple critical domains?”

### New knowledge deduced from the precision nomothetic model

The current PLS model created valuable new knowledge. First, the model demonstrates that depression is made up of many interconnected domains, with lifelong factors such as ROI and mood disorder traits (LT_traits), as well as the severity of the current phenome, being closely interconnected critical events. Notably, the lifetime phenome predicts the severity of the phenome of an acute episode, with ROI and LT_traits explaining around 66% of the variance in the current phenome. In addition, no discriminant validity could be established between ROI and depressive traits, suggesting that both facets are manifestations of the same core, namely the lifetime+Current phenome of depression. These findings are highly significant since they demonstrate that neuroticism and dysthymia are mild variants of the disease “depression” [2, 14]. As previously mentioned, neuroticism and dysthymia can be viewed as prodromes or residual symptoms following an acute episode [14]. Increasing ROI is also substantially associated with these subclinical trait symptoms, suggesting that the relapse-remitting aspect of depression is associated with more chronic subclinical symptoms. As such, the course of depression bears relationships with the course of autoimmune disorders, including multiple sclerosis. While LT_traits may be subclinical symptoms, it may be more appropriate to characterize the intensity of the actual phase of depression as an intensified signal of these traits [14].

The second major finding is that no discriminatory validity could be established between the lifetime (ROI and traits) and current (severity of depression and anxiety scales and decreased HR-QoL) phenome of depression, and that one validated factor could be retrieved from both phenomes. This implies that both the lifetime and the current phenome are manifestations of the same underlying concept, namely depression as a lifelong disease. This again demonstrates that the DSM/ICD binary diagnostic models for MDD are inaccurate. In fact, by combining multiple outcome variables (i.e., lifetime and current phenome characteristics) into a single factor, we integrate numerous bits of data and so increase power and precision (see Introduction). These findings indicate that the outcome of a disorder should not be represented as a binary variable, but rather as a dimensional score based on the disorder’s various facets.

### New clinical precision model of MDMD

We also described how cluster analysis can be used to identify new groups of patients with similar characteristics and identified a cluster of patients with elevated ACEs, ROI, LT_traits, and lifetime and current SI, SA, and SB scores as well as phenome scores. Using various cluster analysis techniques, a similar group of patients was identified in previous research and labelled “major dysmood disorder or MDMD”, whereas the remaining depressed subjects were classified as having “simple dysmood disorder (SDMD)” due to their lower RADAR scores on all features [1,5,7]. We use the term “dysmood” instead of depression, because patients with this condition exhibit more than just depressive symptoms, including anhedonia, hypoesthesia, diurnal variation, ruminations, irritability, anxiety, dysthymia, physiosomatic symptoms, SBs, disabilities, cognitive impairments, and a decreased HR-QoL [1,5,7]. In addition, patients with bipolar disorder can be classified as MDMD and SDMD and, in fact, they do not differ from unipolar depressed MDD patients in any of the model’s features [7]. Hence, from now on, we will refer to depression as MDMD or SDMD, which better describes the condition.

Besides from considering all RADAR scores, the diagnosis of MDMD is also useful for clinical and research purposes. First, it is a more precise model of depression than MDD because it is based on all available data. Second, the introduction of a new label based on a quantitative score of Lifetime+Current phenome features is better suited to remedy the usual mistake made by psychiatrists and psychologists, which is to think in terms of having MDD or not. An even greater difficulty is that, as described in the introduction, SDMD is a heterogeneous condition that frequently includes patients with normal distress responses, such as grief, demoralization, and human sadness (see Introduction). Moreover, the clinical diagnosis MDMD, but not SDMD, is reified as a biological disorder. There are indeed qualitative differences in many AOPs between MDMD and SDMD. Thus, in the present study, we found that the cluster-analytically-derived MDMD cluster is externally validated by increased atherogenicity scores, whereas patients with SDMD occupied an intermediate position between MDMD patients and controls. Previously, we found that MDMD, but not SDMD, is associated with immune activation, activation of the growth factor network, PON1 Q192R variants, lowered PON1 activity, nitrosative stress, oxidative stress, leaky gut, gut dysbiosis, and autoimmunity [5,6,11,14,15,16]. Consequently, MDMD, which is characterized by increased ROI, LT_traits, increased phenome severity and diverse AOPs, is a qualitatively distinct “phenotype and pathway class”. As a result, using MDD (i.e. combining SDMD and MDMD) hinders the discovery of new biomarkers and pathways. Therefore, we believe that future research on depression should recruit controls and MDMD patients, rather than MDD or SDMD patients, and develop new diagnostic biomarkers or tools (combinations of biomarkers using support vector machine or neural networks) to predict MDMD.

Examining the progression of SDMD patients is another important topic for future research. It is likely that a subset of first episode SDMD patients may later exhibit increasing ROI scores, thereby developing MDMD. The genetic and risk biomarkers of ROI will hopefully be able to differentiate the group of SDMD patients into those who will develop more episodes (and thus MDMD) and those who suffer from normal human distress responses.

### Incorporation of atherogenicity in the precision nomothetic model

In this study, we also explained how to incorporate biomarkers into the precision model, and how to compute pathway scores (e.g. PRO/ANTI_AI) that reflect the degree of atherogenicity. Our model showed us that atherogenicity is strongly associated with all features of dysmood disorder, namely ROI, lifetime and current SBs, and severity of the current phenome, including lowered HR-QoL and the subclinical traits, which may precede acute episodes of dysmood disorders. In this respect, we should emphasize that previous research in dysmood disorders, discovered that lowered activity of the PON1 enzyme, partly due to alterations in Q192R PON1 variants, is associated with increasing ROI and phenome scores [18]. After release from the liver into the serum, the PON1 molecule binds to HDLc to form a HDLc-PON1-ApoA complex which determines RCT and, thus, the clearance of free cholesterol from the tissues and increased atherogenicity [13,18,20,21,23,36]. As a result, Q192R variants, which are predictors of ROI and the phenome of MDMD, play a role in increased atherogenicity in MDMD.

In addition, as previously explained, the HDLc-PON1-ApoA complex is a major anti-inflammatory and antioxidative pathway that protects against antimicrobial activity and increased LPS load and subsequent Toll Like Receptor-4 stimulation [13,18,20,21,23,36]. As a result, impairments in RCT may drive atherogenicity and immune and oxidative pathways, as well as bacterial translocation, which together may determine many aspects of the MDMD phenome [6,11,16,17]. Consequently, atherogenicity and decreased RCT are components of the “microimmuneoxysome” pathophysiology of mood disorders [13].

It is important to note that ACEs explained a larger part (around 22%) of the variance in the Lifetime+Current phenome of MDMD. Such findings extend those of previous papers that ACEs predict ROI, neuroticism, severity of illness, suicidal behaviors, and lowered HR- QoL [13, 14]. Moreover, our results show that part of the effects of ACEs on the phenome of depression are mediated by increased atherogenicity. Previously, we established that ACEs are associated with immune activation, PON1 enzymatic activity, activation of the growth factor network and a specific gut enterotype [5,6,11,15,16,17]. This suggests that genetic Q192R variants and ACEs both affect the same AOPs that lead to MDMD. Moreover, a previous paper showed that an interaction between ACE x the Q192R PON1 genetic variant is a significant predictor of ROI and the phenome [9, 10].

### RADAR scores in clinical practice

Evidently, in research and clinical settings, the various RADAR scores and MDMD diagnosis should be utilized as outcome variables. However, most scientists and clinicians do not comprehend the ML techniques employed in this study, namely factor and multiple regression analysis combined with PLS-SEM. As a result, they will be hesitant to embrace the new method and will continue to implement the incorrect binary gold standards. Nevertheless, the RADAR scores are extremely simple to calculate. Using the theoretical framework depicted in Figure 1 and PLS software, any statistician should be able to compute a PLS model and derive the latent variable scores. As a result, it is quite simple to compute the models, and data from new patients can be automatically projected into the model and utilized to generate the RADAR graph.

The addition of atherogenicity indices to the RADAR graph gives extra information and indicates tailored medication targets for patients. Thus, increased atherogenicity scores or decreased RCT scores suggest that the patient should be treated with treatments that increase HDLc and PON1 and decrease atherogenicity, such as omega-3 and omega-9 (oleic acid) polyunsaturated fatty acids, niacin (vitamin B3), exercise, weight loss, and statins combined with coenzyme Q10 [18,36,37].

## Limitations

Clearly, other microimmuneoxysome pathways should be added to the RADAR graph and be used as therapeutic targets, such as oxidative stress, lipid peroxidation, immunological activation, activation of the growth factor network, JAK-STAT pathway, nitrosative stress, and autoimmunity to neoantigens [5,6,11,15,16,17].

## Conclusions

Our results indicate that the outcome of depression should not be represented as a binary diagnostic variable, but rather as multidimensional scores based on biomarkers, ROI, and other lifetime and current phenome scores. One factor could be extracted from a) the lifetime phenome of depression, which included ROI and mood disorder traits, and b) the acute phase phenome of depression. Increased atherogenicity, neglect, and sexual abuse explained 55.7% of the variance in the Lifetime+Current phenome factor. Cluster analysis identified a cluster of patients with MDMD, a qualitatively distinct pathway-class, which was externally validated by elevated atherogenicity and defined by elevated scores for all clinical features.

Our new approach will considerably improve the development of biomarker research. Instead of association studies that examine the links between risk biomarkers and a flawed binary MDD diagnosis or diagnostic biomarkers that confirm the MDD binary diagnosis, our method allows one to develop a multitude of different biomarker tools. First, new patient data may be projected into our PLS model, and a customized RADAR graph can be readily generated, providing appropriate clinical monitoring and biomarker monitoring of the condition. Second, our method allows future research to compute a) new risk or susceptibility biomarkers of MDMD, ROI, suicidal behaviors; mood disorder traits, and HR-Qol; b) new screening and detecting biomarkers as well as diagnostic biomarker panels of MDMD, suicidal behaviors; ROI and traits; c) new prognostic molecular biomarkers of ROI, suicidal behaviors, and the progression of subclinical traits and disease; d) new stratification biomarkers to define the groups of MDMD patients who will respond to new treatments; e) new efficacy or outcome biomarker tools which help to detect an early response in the treatment of ROI, suicidal behaviors and illness severity; and f) new predictive biomarker tools that will guide personalized treatments of MDMD, recurrence of illness or suicidal behaviors.

## Author**’**s contributions

Conceptualization and study design: MM and KJ; first draft writing: MM; editing: all authors; recruitment of patients: KJ; CT and PB; statistical analysis: MM. All authors approved the final version of the manuscript.

## Ethics approval and consent to participate

The research project (#445/63) was approved by the Institutional Review Board of Chulalongkorn University’s institutional ethics board. All patients and controls gave written informed consent prior to participation in the study.

## Data Availability Statement

The dataset that was made and/or analysed during this study will be available from the corresponding author (MM) once it has been fully used by the authors and a reasonable request has been made.

---

**ESF, Table 1.** Demographic and clinical data of the major depression patients (MDD) and healthy controls (HC) included in the present study. Results are shown as mean ±SD. F: results of analysis of variance; X²: analysis of contingency tables, FFHT: Fisher-Freeman-Halton Exact Test; KWT: Kruskal-Wallis test. BMI: body mass index; TUD: Tobacco use disorder; BDI: Beck Depression Inventory; HAMD: Hamilton Depression Rating Scale; STAI: State and Trait Anxiety Inventory; PC: principal component; SB: suicidal behaviors, that is ideation (SI) and attempts (SA); T2DM: type 2 diabetes mellitus; PC HAMD + BDI + STAI: first PC score extracted from these three rating scales; PC HAMD + BDI + STAI + Current SB: first PC score extracted from these three rating scales and suicidal behaviors; HR_QoL: health-related quality of life, T2DM: type 2 diabetes mellitus. Adapted from Jirakran et al., 2023 [14]

| **Variables** | **HC (n=67)** | **MDD (n=66)** | **F/X<sup>2</sup>/FEPT** | **df** | **p** |
| --- | --- | --- | --- | --- | --- |
| Sex (Male / Female) | 9/58 | 18/48 | 3.94 | 1 | 0.055 |
| Age (years) | 37.9 (9.2) | 36.9 (11.5) | 0.263 | 1/131 | 0.609 |
| Education (years) | 14.1 (3.4) | 14.8 (2.8) | 1.72 | 1/131 | 0.193 |
| Income (baht/month) | 21500 (7269) | 23770 (19203) | 0.817 | 1/131 | 0.368 |
| Single/married/widower/divorced | 35/26/3/3 | 36/22/4/4 | FHHET |  | 0.877 |
| Employed / unemployed | 66/1 | 47/19 | 19.388 | 1 | <0.001 |
| TUD (No/Yes) | 62/5 | 54/12 | 3.43 | 1 | 0.074 |
| BMI (kg/m<sup>2</sup>) | 27.4 (6.1) | 26.9 (5.5) | 0.266 | 1/130 | 0.607 |
| Waist Circumference (cm) | 88.8 (14.9) | 89.7 (15.0) | 0.142 | 1/130 | 0.707 |
| Systolic Blood Pressure (mmHg) | 127.9 (15.3) | 129.9 (17.8) | 0.475 | 1/131 | 0.492 |
| Diastolic Blood Pressure (mmHg) | 79.2 (11.3) | 77.9 (12.4) | 0.420 | 1/131 | 0.518 |
| Metabolic syndrome (MetS) (No/Yes) | 34/33 | 35/31 | 0.069 | 1 | 0.863 |
| Neuroticism | 18.3 (5.0) | 27.2 (6.4) | 80.332 | 1/131 | <0.001 |
| Total BDI | 5.6 (6.5) | 26.9 (13.8) | 129.518 | 1/131 | <0.001 |
| Total HAMD | 2.3 (3.0) | 18.1 (6.4) | 330.943 | 1/131 | <0.001 |
| Total STAI | 38.2 (7.8) | 51.9 (10.4) | 73.623 | 1/131 | <0.001 |
| PC HAMD + BDI + STAI | -0.758 (0.421) | 0.770 (0.810) | 187.443 | 1/131 | <0.001 |
| PC HAMD + BDI + STAI + Current SB | -0.754 (0.352) | 0.765 (0.850) | 182.045 | 1/131 | <0.001 |

| Variables | HC (n=67) | MDD (n=66) | F/X<sup>2</sup>/FEPT | df | p |
| --- | --- | --- | --- | --- | --- |
| Current suicidal ideation (SI) | -0.489 (0.0) | 0.493 (1.248) | KWT | - | <0.001 |
| Current suicidal attempts (SA) | -0.245 (0.0) | 0.256 (1.390) | KWT | - | 0.004 |
| Current Suicidal behaviors (SI and SA) | -0.420 (0.0) | 0.426 (1.290) | KWT | - | <0.001 |
| HR_QoL, Physical domain | 26.9 (3.4) | 21.5 (4.5) | 61.92 | 1/131 | <0.001 |
| HR_QoL, mental health | 23.0 (3.6) | 17.3 (5.6) | 46.65 | 1/131 | <0.001 |
| HR_QoL, social | 11.6 (2.0) | 9.2 (2.5) | 37.02 | 1/131 | <0.001 |
| HR_QoL, environment | 28.4 (4.9) | 25.4 (5.8) | 10.32 | 1/131 | 0.002 |
| Previous Covid-19 infection (No/Yes) | 62/5 | 62/4 | 0.104 | 1 | 1.000 |
| T2DM (No/Yes) | 61/6 | 61/5 | 0.083 | 1 | 1.000 |
| Dyslipidemia (No/Yes) | 58/9 | 55/11 | 0.272 | 1 | 0.635 |
| Drugs targeting MetS-related conditions<br>(No/Yes) | 50/17 | 52/14 | 0.322 | 1 | 0.682 |

Results are shown as mean ± SD. F: results of analysis of variance; X<sup>2</sup>: analysis of contingency tables, FFHT: Fisher-Freeman-Halton Exact Test; KWT: Kruskal-Wallis test.
**ESF, Table 2.** Biomarker data of the major depression patients (MDD) and healthy controls (HC) without metabolic syndrome.

**ESF, Table 2.** Biomarker data of the major depression patients (MDD) and healthy controls (HC) without metabolic syndrome. Results are shown as estimated marginal means (SEM) after univariate GLM analyses with age, sex, body mass index and waist circumference as covariates; * Processed in Log transformation; HDLc: high density lipoprotein cholesterol; LDLc: low density lipoprotein cholesterol; PRO_AI: composite based on LDLc, TG, ApoB, and FC; ANTI_AI: composite based on HDLc, ApoA and CER; PRO/ANTI_AI: PRO_AI versus ANTI_AI ratio Adapted from Jirakran et al., 2023 [14]

| Variables | HC (n=34) | MDD (n=35) | F | df | p |
| --- | --- | --- | --- | --- | --- |
| Free Cholesterol (FC) (mg/dL) | 51.7 (3.2) | 62.0 (3.2) | 5.00 | 1/62 | 0.029 |
| Cholesteryl esters (mg/dL) | 138.3 (5.6) | 140.8 (5.6) | 0.10 | 1/62 | 0.756 |
| Cholesterol esterification rate (CER) | 72.8 (1.5) | 69.2 (1.5) | 2.72 | 1/62 | 0.104 |
| HDLc (mg/dL) | 64.3 (2.4) | 56.8 (2.4) | 4.83 | 1/62 | 0.032 |
| Triglycerides (TG) (mg/dL) * | 84.1 (8.9) | 123.1 (8.9) | 9.71 | 1/62 | 0.003 |
| LDLc (mg/dL) | 118.2 (6.2) | 132.3 (6.2) | 2.52 | 1/62 | 0.118 |
| Apolipoprotein A (ApoA) (mg/mL) | 149.8 (5.0) | 135.2 (5.0) | 4.14 | 1/62 | 0.046 |
| ApoB (mg/mL) | 80.6 (3.6) | 91.5 (3.6) | 4.39 | 1/62 | 0.040 |
| ApoB/ApoA ratio * | 0.56 (0.03) | 0.70 (0.03) | 10.34 | 1/62 | 0.002 |
| TG/HDLc | 1.37 (0.22) | 2.39 (0.22) | 10.75 | 1/62 | 0.002 |
| PRO_AI (z scores) | -0.308 (0.170) | 0.325 (0.170) | 6.73 | 1/62 | 0.012 |
| ANTI_AI (z scores) | 0.308 (0.165) | -0.286 (0.165) | 6.29 | 1/62 | 0.015 |
| PRO/ANTI_AI (z scores) | -0.392 (0.163) | 0.384 (0.163) | 11.00 | 1/62 | 0.002 |

**ESF, Figure 1.**
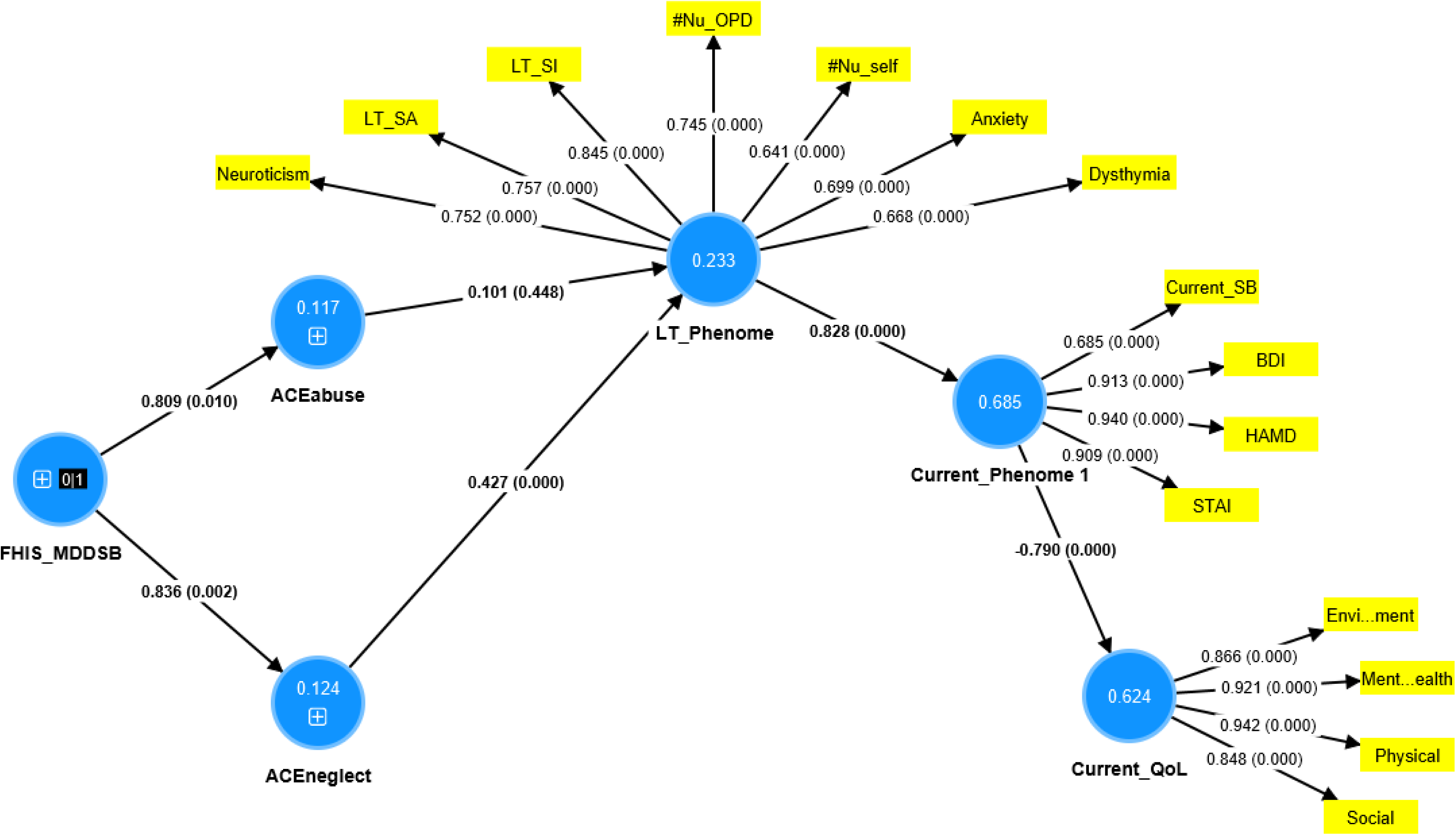
A second Partial Least Squares model. Current_QoL: latent vector (LV) extracted from 4 health-related quality of Life (QoL) domains; Current_phenome: LV extracted from current suicidal behaviors (Current_SB), Hamilton Depression Rating Scale (HAMD), Beck Depression Inventory (BDI), and Spielberger State and Trait Anxiety; LT_phenome: a LV extracted from lifetime suicidal ideation (LT_SI) and attempts (LT_SA), and number of episodes, either according to the outpatient files (#Nu_OPD) or self-declared (#Nu_self), and mood disorder traits; ACE: adverse childhood experiences, either abuse or neglect; FHIS_MDDSB: family history in first-degree relatives of major depression and suicidal attempts.

**ESF, Figure 2.**
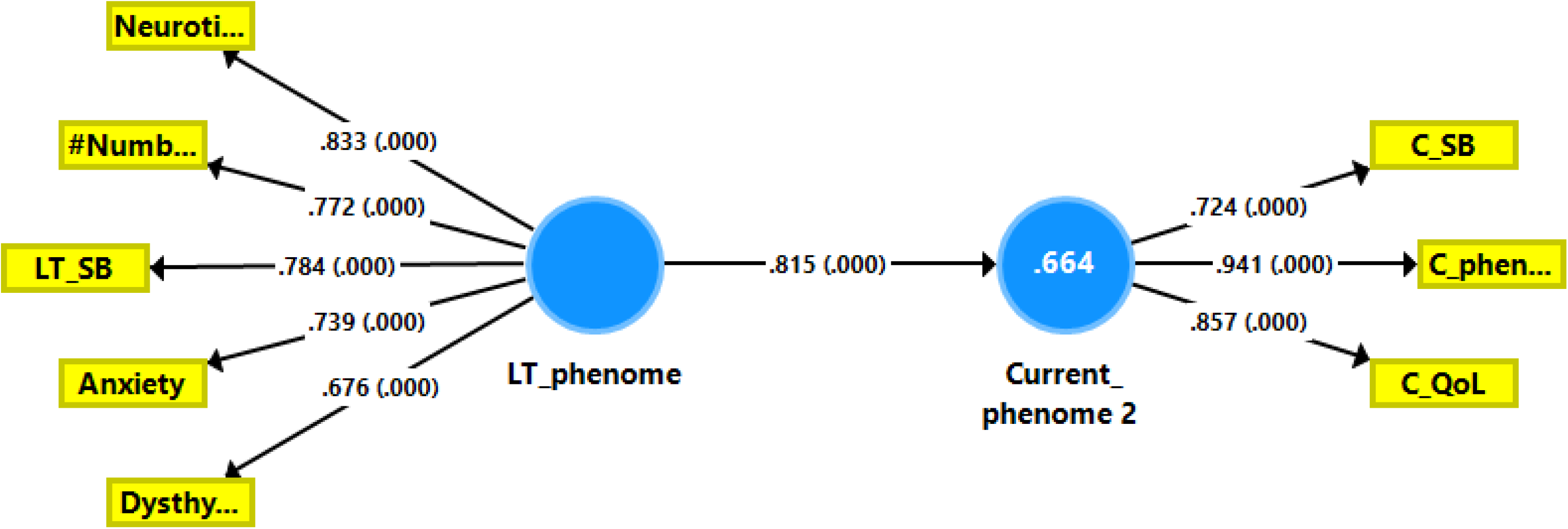
A third Partial Least Squares model. Current_phenome 2: a latent vector (LV) extracted from current suicidal behaviors (C_SB), C_phenome (a LV extracted from the Hamilton Depression Rating Scale, Beck Depression Inventory, and Spielberger State and Trait Anxiety scores) and C_QoL (a LV extracted from the 4 quality of life domains); LT_phenome: a LV extracted from lifetime (LT) suicidal ideation (LT_SI) and attempts (LT_SA), and number of episodes (#Numb), and mood disorder traits.

